# Time varying association between deprivation, ethnicity and SARS-CoV-2 infections in England: a space-time study

**DOI:** 10.1101/2021.11.09.21266054

**Authors:** Tullia Padellini, Radka Jersakova, Peter J. Diggle, Chris Holmes, Ruairidh E. King, Brieuc C. L. Lehmann, Ann-Marie Mallon, George Nicholson, Sylvia Richardson, Marta Blangiardo

**Affiliations:** MRC Centre for Environment and Health, Dept of Epidemiology and Biostatistics, Imperial College London; The Alan Turing Institute, London, UK; CHICAS, Lancaster Medical School, Lancaster University, UK; University of Oxford, UK; MRC Harwell Institute, Harwell, UK; MRC Biostatistics Unit, University of Cambridge, UK

## Abstract

**Background:** Ethnically diverse and socio-economically deprived communities have been differentially affected by the COVID-19 pandemic in the UK.

**Method:** Using a multilevel regression model we assess the time-varying association between SARS-CoV-2 infections and areal level deprivation and ethnicity. We separately consider weekly test positivity rate (number of positive tests over the total number of tests) and estimated unbiased prevalence (proportion of individuals in the population who would test positive) at the Lower Tier Local Authority (LTLA) level. The model also adjusts for age, urbanicity, vaccine uptake and spatio-temporal correlation structure.

**Findings:** Comparing the least deprived and predominantly White areas with most deprived and predominantly non-White areas over the whole study period, the weekly positivity rate increases by 13% from 2·97% to 3·35%. Similarly, prevalence increases by 10% from 0·37% to 0·41%. Deprivation has a stronger effect until October 2020, while the effect of ethnicity becomes slightly more pronounced at the peak of the second wave and then again in May-June 2021. Not all BAME groups were equally affected: in the second wave of the pandemic, LTLAs with large South Asian populations were the most affected, whereas areas with large Black populations did not show increased values for either outcome during the entire period under analysis.

**Interpretation:** At the area level, IMD and BAME% are both associated with an increased COVID-19 burden in terms of prevalence (disease spread) and test positivity (disease monitoring), and the strength of association varies over the course of the pandemic. The consistency of results across the two outcome measures suggests that community level characteristics such as deprivation and ethnicity have a differential impact on disease exposure or susceptibility rather than testing access and habits.

**Fundings:** EPSRC, MRC, The Alan Turing Institute, NIH, UKHSA, DHSC, NIHR

## Introduction

The differential impact of COVID-19 on minorities has reportedly varied over the course of the pandemic. An early review of the literature by Public Health England (1) concluded there is evidence of higher test positivity rates, defined as the percentage of processed SARS-CoV-2 swab tests that are positive, for people identifying as Black. Later studies reported an increased test positivity rate and mortality for the Black, Asian and Minority Ethnic (BAME) population in the first wave of the pandemic in England (2) and Wales (3). Mathur and colleagues additionally found that between September and December 2020 the increased risk of testing positive disappeared and there was a reduced risk of death for Black and mixed ethnicity groups but not for South Asians (2). Morales and Ali (2021) suggested that this change might be due to interventions that occurred between the two waves (e.g. better access to testing, targeted public communication and reduction of occupational risks), but stressed the importance of additional studies to better disentangle the effect of ethnicity from that of other variables (4).

Socio-economic deprivation is another key factor that contributes to health inequalities. People living in the most deprived areas in England and Wales were reportedly twice as likely to die from COVID-19 in the first wave (5). A Europe-wide study tested a range of socio-demographic characteristics at country level and found that income was the best predictor of COVID-19 cases between December 2019 and April 2020 (6). As socio-economic deprivation correlates with ethnicity^1^, it is necessary to consider both dimensions simultaneously to accurately assess their impact on COVID-19 burden. Socio-economic and ethnic inequalities in England have become more pronounced during the pandemic and have contributed to unequal death tolls across communities (7). Rose and colleagues found an association between socio-economic deprivation and mortality in England at Upper Tier Local Authority level in the first wave, after adjusting for ethnicity and other socio-demographic factors such as population density and age (8). A recent study by Lo et al. reported that ethnic minorities were at a higher risk of testing positive in the UK and the US in 2020 and some of this increase could be explained by these communities living in more deprived areas (9).

A full detailed analysis of whether and how the role of social and ethnic inequalities on SARS-CoV-2 infections varied over time is still missing from the literature. This is especially true for understanding their combined effect. Most literature to date covers March to December 2020 and has focused either on mortality or test positivity rates, with the latter indicator being affected not only by the actual number of infections but also by testing capacity and strategies. In this study we focus on SARS-CoV-2 infections in England and evaluate how the effects of area levell socio-economic deprivation and ethnic composition evolved over the course of the pandemic (from 1^st^ June 2020 to 19^th^ September 2021). A key novelty of our analysis is that we consider test positivity rate as well as a new derived measure of prevalence proportion which corrects for the ascertainment bias of the tested population, enabling us to assess the impact of inequalities both on observed disease surveillance indicators and on the estimated underlying infection rate.

## Methods

We examine the effects of inequalities on two outcome metrics: test positivity rate and unbiased prevalence proportion. We analyse outcomes as weekly summaries at Lower Tier Local Authority (LTLA) level, the finest spatial resolution at which the UK government monitors epidemic indicators and implements local interventions against COVID-19. Due to data sparsity we remove Isles of Scilly, and, in accordance with government reporting of COVID-19 cases, we combine the Boroughs of Hackney and City of London, leaving 312 LTLAs in the analysis. Aggregating data by week mitigates day-of-week effects in administering, processing and reporting the outcomes of interest. Our analysis covers the period from the 1st of June 2020 to the 19^th^ of September 2021. We exclude data from the first wave of the pandemic (March - May 2020) since community testing was not widely available. Data is stratified by age (5-12, 13-17, 18-24, 25-34, 35-44, 45-54, 55-64, 65+ years old); we do not consider 0-4 years old, as prevalence estimates are not available for this age group.

### Outcomes

Pillar 2^2^ tests provide a measure of transmission in the community and have been one of the key metrics used in decision-making throughout the pandemic. We focus on test positivity rate, which is the number of positive tests as a proportion of the total number of processed tests. We retrieve weekly test data from the PHE Second Generation Surveillance System (SGSS) database, which contains both swab tests (PCR/qrtPCR, LAMPore) and Lateral Flow Testing (LFT) kits at the individual level. We restrict the main analysis to PCR tests only, and report results for combined PCR+LFT data as a sensitivity analysis.

The number of positive tests provides some indication of the epidemic dynamics, but suffers from ascertainment bias: in view of the Test and Trace recommendations, individuals are more likely to get tested if they present symptoms or are in specific working categories (e.g healthcare). As a result, test positivity may be better suited as a measure of disease monitoring than disease spread. We therefore separately analyse weekly point prevalence proportion, defined as the proportion of infected individuals in the population who would be PCR-positive if tested. Debiased prevalence estimates are obtained as output of the methodology developed in Nicholson et al. (10) which combines PCR Pillar 2 testing data with randomised surveillance data from the Real-time Assessment of Community Transmission (REACT)^3^ study (11).

### Exposure variables

We consider two community-level characteristics: (i) social deprivation, measured through the 2019 Index of Multiple Deprivation (IMD)^4^ and (ii) ethnic diversity, defined as the proportion of BAME (Black, Asian and Minority Ethnic) population in each geographical area as reported in the 2011 Census. The distribution of these two covariates by LTLA is presented in Figure 1 of Supplementary material. We also repeat the analysis disaggregating BAME into Black, South Asian (those identifying as Pakistani, Indian or Bangladeshi) and Other BAME (people not identifying as White, Black or South Asian), and include all three as covariates. Figure 2 in Supplementary material presents the proportion of people from a South Asian, Black and Other BAME background in each LTLA and shows the strongly spatially clustered nature of this variable.

**Figure 1:**
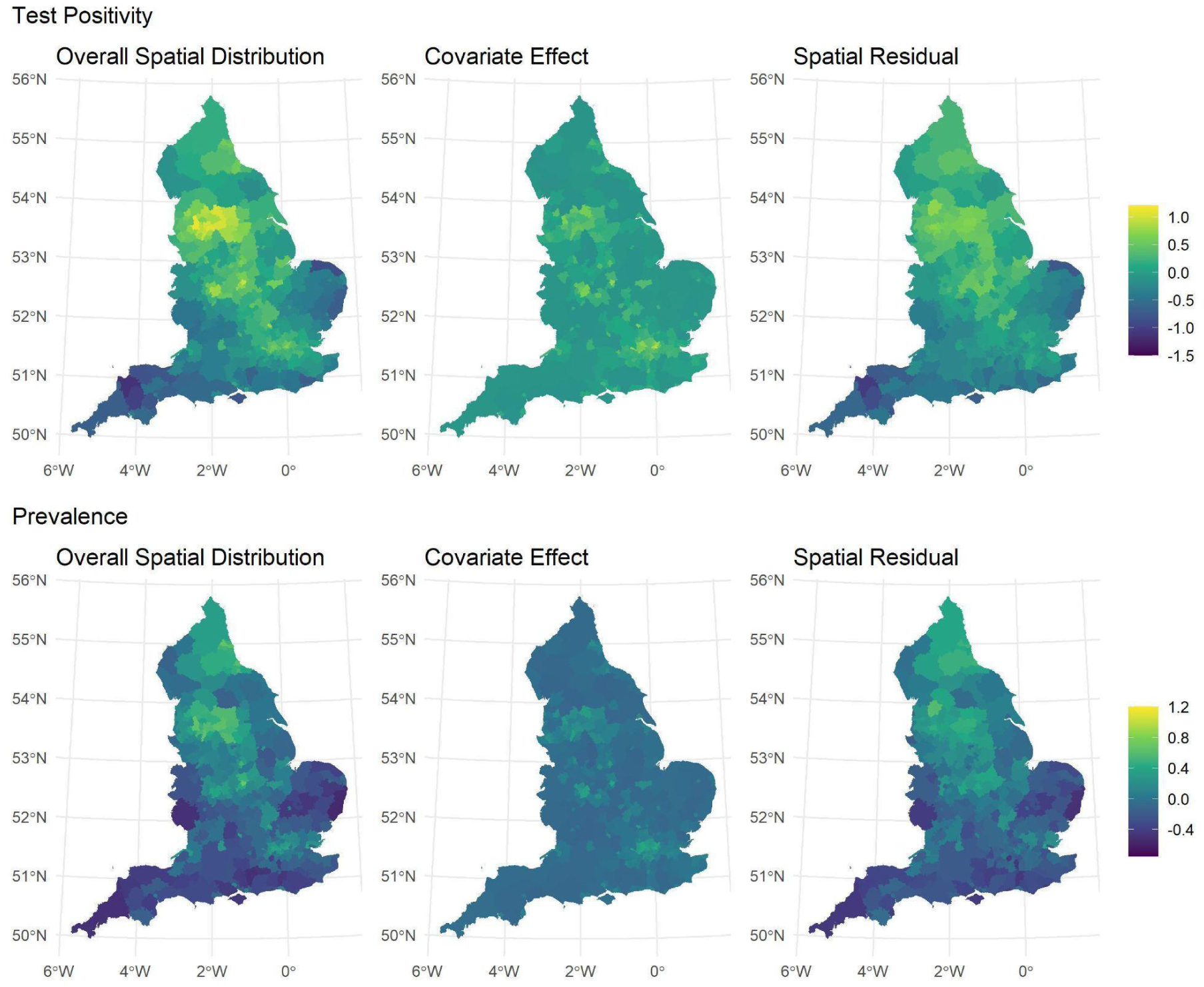
Spatial distributions of the median test positivity (top panel) and debiased prevalence (bottom panel) over the entire study period outputted by the model, presented on the logit scale. The overall spatial distribution (left column) is disaggregated into two components, representing what is explained by social deprivation, ethnic composition and confounders (centre column) and the unobserved spatial residuals (right column).

**Figure 2:**
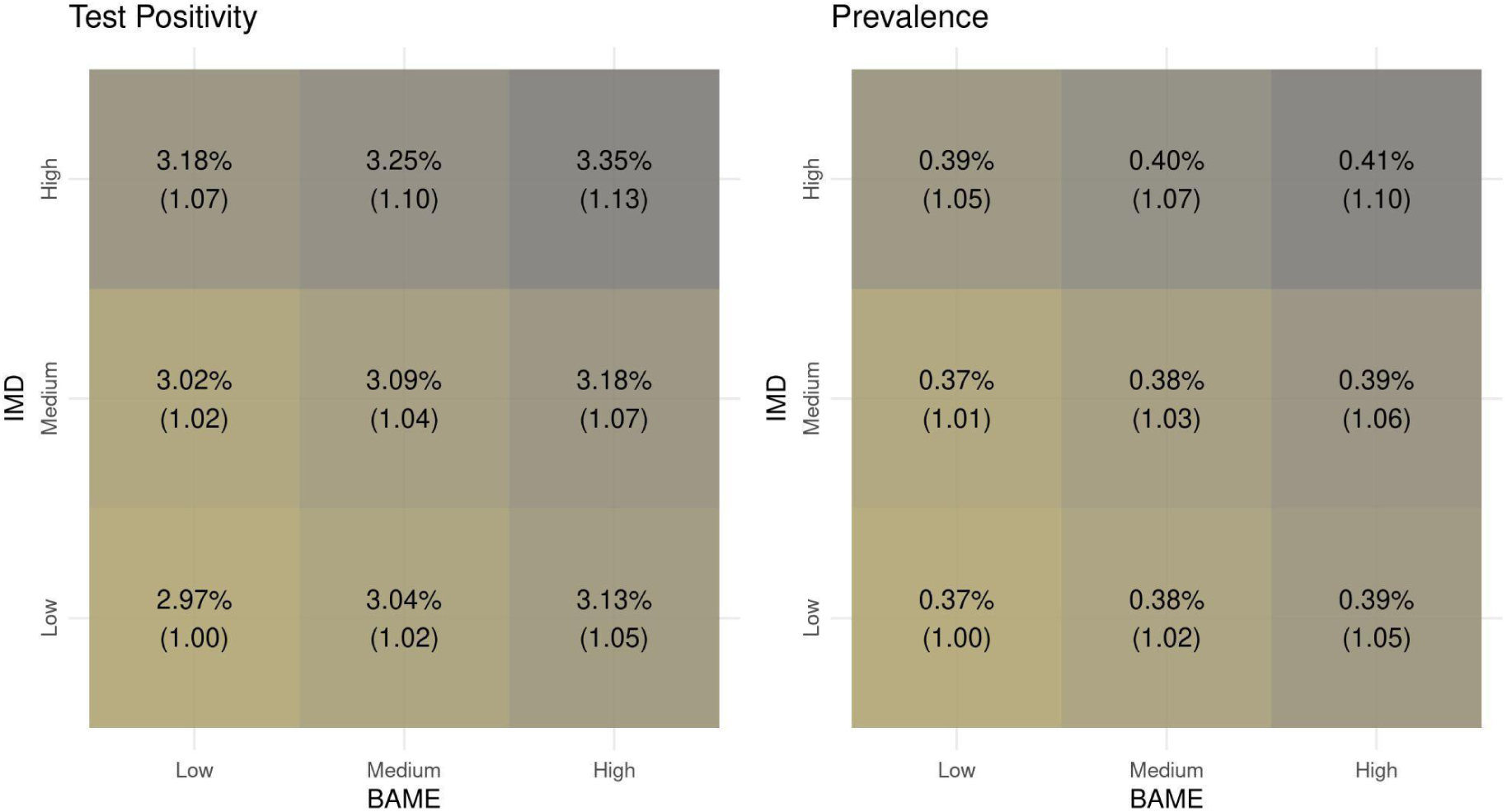
Test positivity (left) and debiased prevalence (right) for profiles of ethnicity (all BAME) and deprivation. Each tile represents the average weekly outcome (test positivity or debiased prevalence) over the entire period of analysis, obtained as output of the model including ethnicity, IMD, confounders and the spatio-temporal correlation structure. In parenthesis we report the relative change in the outcome between each profile and the reference, defined as low deprivation and low BAME population.

To illustrate the combined effect of ethnicity and deprivation, we define profiles based on the distribution of the IMD score and the percentage of BAME population across LTLAs. We consider as a representative *low* value the first quartile, that is, the value that divides the bottom 25% from the remainder of the distribution of the variable. Conversely, we consider a representative *high* value to be the one dividing the bottom 75% from the remaining 25%. Finally we consider the median as a representative *medium* value.

### Confounders

As confounders we consider the level of urbanicity and the percentage of the population that is fully vaccinated. LTLAs are divided into “ Predominantly Rural”, “ Predominantly Urban” and “ Urban with Significant Rural” following the 2011 Rural-Urban Classification of Local Authority Districts in England.^5^ Vaccination data is retrieved from PHE and contains counts of vaccinations with Vaxzevria (Oxford University/Astrazeneca), Comirnaty (Pfizer/BioNTech), COVID-19 Vaccine Moderna (Moderna) and Janssen (Johnson & Johnson). We define as “ fully vaccinated” individuals who received the second vaccine dose (first dose for Janssen) at least two weeks prior. Additionally, in a sensitivity analysis we include an indicator of non-pharmaceutical interventions (NPIs), which captures national lockdowns (5^th^ November - 1^st^ December 2020 and 6^th^ January - 28^th^ March 2021) and LTLAs placed in tier 3 during the local tier system implemented in Autumn 2020^6^.

### Statistical Analysis

We evaluate the association between the two community-level characteristics and the monitoring and spread of COVID-19 using a Bayesian spatio-temporal regression model. This is a common approach designed to overcome high variability in estimates driven by the small numbers of cases at high spatio-temporal resolution (12) (13) (14). For each outcome (in a given LTLA and week) we model the logit-transformed test positivity rate or prevalence proportion as a function of the area-level ethnicity groups and deprivation, with time-varying coefficients. We account for confounding effects of area structure (urban/rural status), policy intervention (vaccination rollout) and age effects on disease susceptibility. After accounting for the effect of ethnicity, deprivation and confounders, the residual variability is decomposed into a spatially structured component indexed by the LTLAs, a temporal trend component indexed by weeks and unstructured independent spatio-temporal residuals. We follow (15) to capture dependence in space, using common boundaries to define spatial contiguity. We model the temporal trend component flexibly as a second order random walk, whereby each week’s value depends directly on the values in the two previous weeks.

Analyses were performed in R using R-INLA (16). A full specification of the model structure can be found in Section 1 of the Supplementary Material. We display maps and time plots of posterior medians for the spatial and temporal trends respectively. We report the posterior median of the outcomes for profiles of ethnicity and deprivation, as well as 95% credible intervals (CI) for the odds ratio (OR) quantifying the effect of the two covariates over the entire period and how it changed in time. For summary of all analyses see Table 1 of the Supplementary Material. The analysis code is provided at https://github.com/alan-turing-institute/jbc-turing-rss-equality.

### Data statement

Test and vaccination data aggregated by age class, LTLA and week are held by PHE and were available to us through the partnership with UKHSA. We are not allowed to share these with third parties. Anyone interested in accessing the data should contact PHE.

## Results

### Overall spatial distribution and temporal trends

The median of logit-transformed test positivity and debiased prevalence outputted by the model is shown in Figure 1 (left column) and depicts the spatial contrasts for the whole period. As shown in the central panel of Figure 1, social deprivation, ethnic composition and the confounders partially explain the higher values of both outcomes in large cities, like London, Birmingham, and the area around Manchester, Leeds and Sheffield. Altogether, exposures and confounders account for approximately 24% of the overall spatial distribution of test positivity and 20% of debiased prevalence, reaffirming the need for including spatially structured residuals in the model (Figure 1, right panel).

The median of the temporal component in the model captures the overall time trend for the two outcomes, shown in Figure 3 in Supplementary material. Both test positivity rate and debiased prevalence show three peaks during the period under analysis. The first two peaks, in November 2020 and December 2020, were curbed by the introduction of a national lockdown (shown in grey). The third, highest, peak is in July-August 2021. When including LFT in the analysis the summer peak is lower than the two winter ones (Figure 4 in Supplementary material, top panel). This might potentially reflect the change in testing policy: LFT became more widespread as businesses were encouraged to sign up for a testing scheme for their staff, increasing the denominator of the positivity rate. The summer peak is also somewhat less pronounced when removing vaccine uptake from the confounders; in this case the temporal random effect implicitly accounts for the fact that higher vaccination rates lead to a reduction in infections (Figure 4 in Supplementary material, central panel). Finally, the time pattern does not change when we include a lockdown indicator, suggesting that the temporal component already captures much of this effect (Figure 4 in Supplementary material, bottom panel).

**Figure 3:**
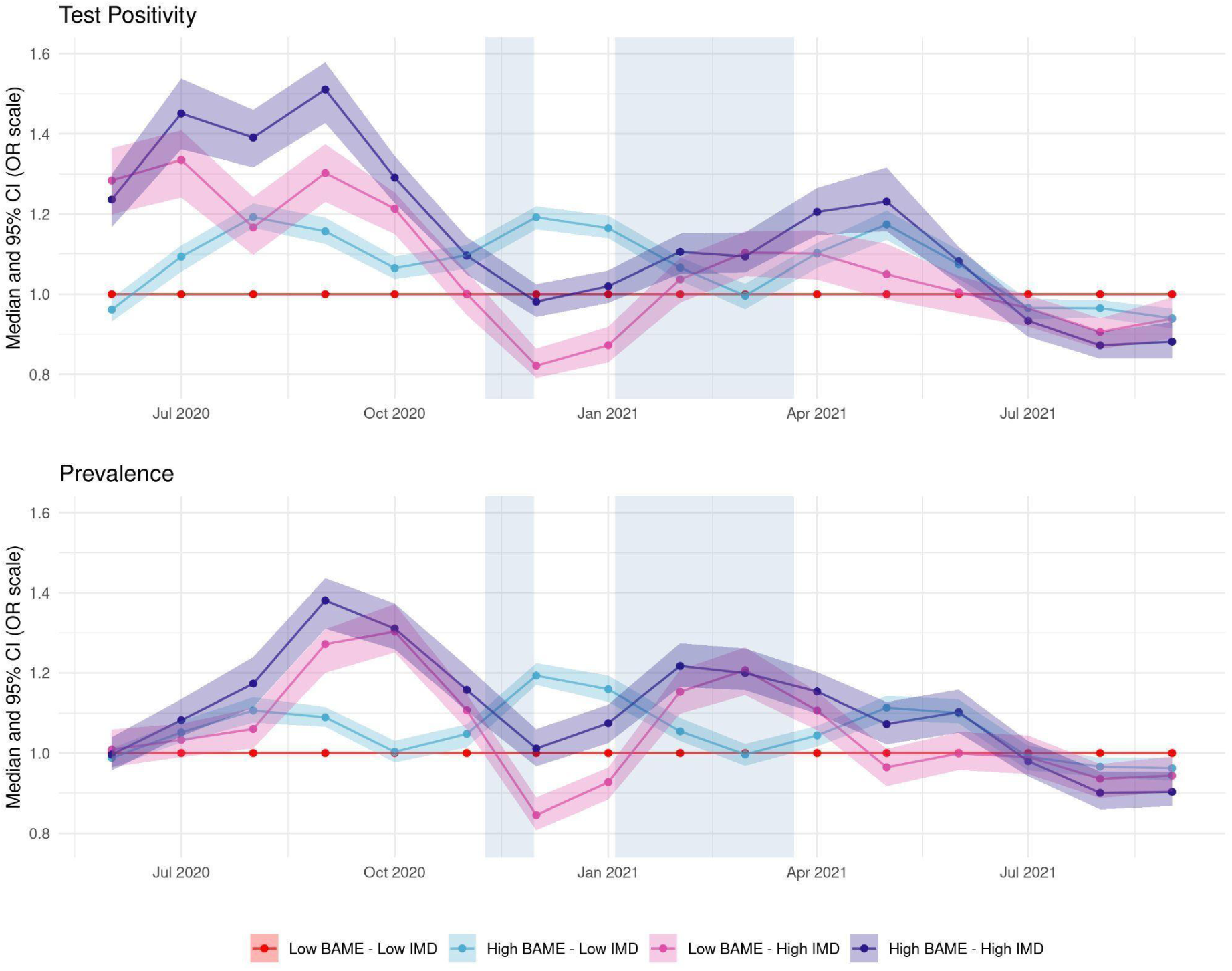
Time-varying test positivity (top) and debiased prevalence (bottom) for profiles of ethnicity (all BAME) and deprivation. Each line represents the monthly median odds ratio for the profile relative to a population low in deprivation and BAME, while the shaded area around it represents the corresponding 95% credible interval. The median odds ratio and its uncertainty are outputs of the model after adjusting for confounders and the spatio-temporal correlation structure. Periods of national lockdown are marked in grey.

**Figure 4:**
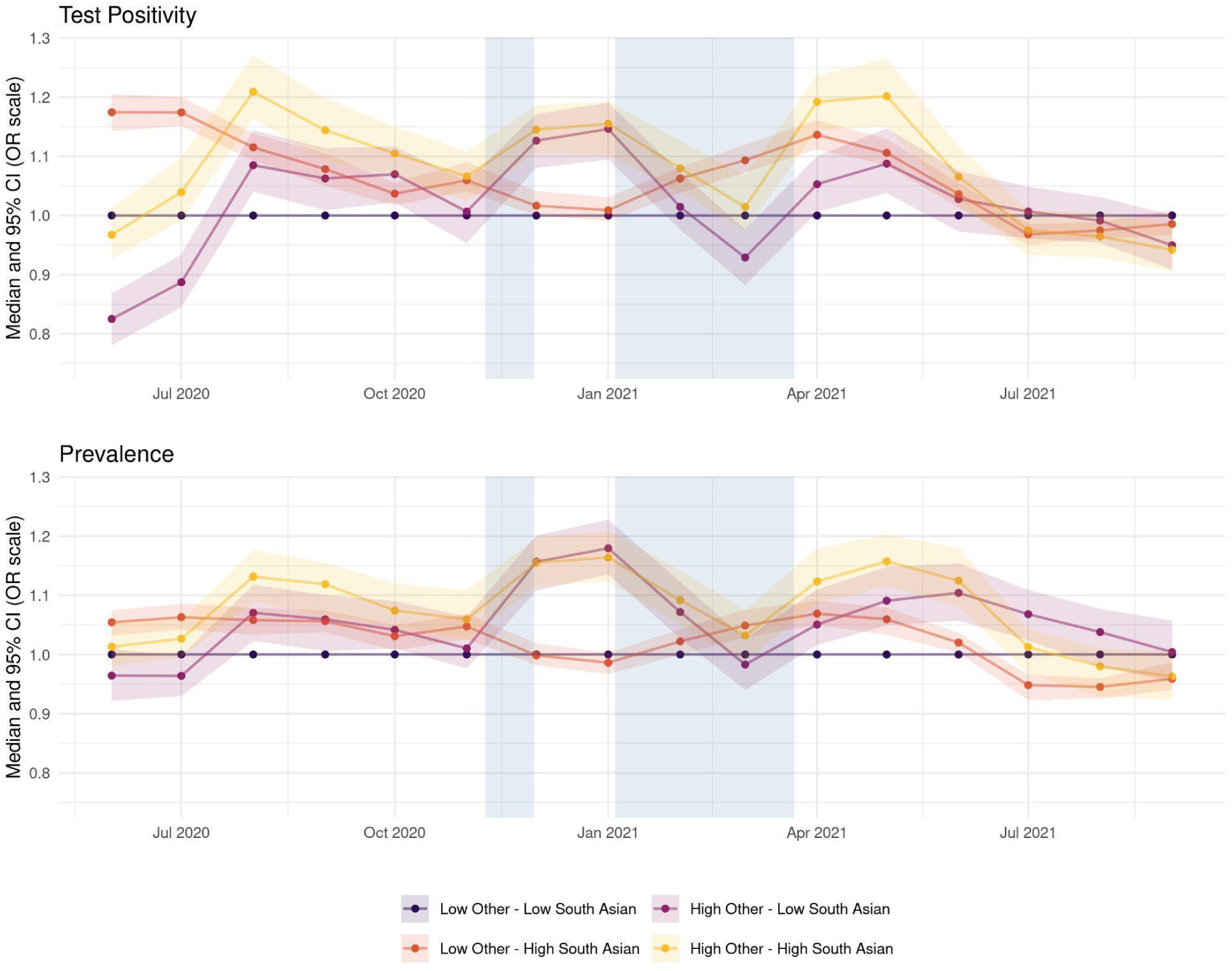
Time-varying test positivity (top) and debiased prevalence (bottom) for profiles of ethnicity disaggregated by BAME subgroups (South Asian, Other). Each line represents the monthly median odds ratio for that profile relative to a population with a low percentage of all BAME subgroups, while the shaded area around it represents the corresponding 95% credible interval. The median odds ratio and its uncertainty are outputs of the model after adjusting for IMD, confounders and the spatio-temporal correlation structure. Periods of national lockdown are marked in grey.

### Overall effects of deprivation and ethnicity

The effects of the variables included in the model are reported in Figure 5 of Supplementary material as odds ratio (OR), i.e. change in the outcome (represented on odds scale) associated with a 1 standard deviation (SD) increase in one of the exposure variables (as these are standardised). One SD corresponds to a 7·9 point increase in the IMD score and to a 13% increase in the aggregated BAME population. For analyses with disaggregated BAME subgroups, 1 SD corresponds to an increase of 4·4% for the Black subgroup, 6·3% for South Asians, and 4·3% for Other BAME.

An increase in South Asian population proportion corresponds to higher values of both outcomes: OR is 1·1 (95% CI 1·07-1·13) for test positivity and 1·04 (95% CI 1·01-1·06) for prevalence. The associations between Other BAME population and the two outcomes are characterised by larger uncertainty, with OR of 1·02 (95% CI 0·98-1·07) for test positivity and 1·06 (95% CI 1·02-1·10) for prevalence. There is not enough evidence to suggest that an increase in Black population is associated with an increase in either test positivity, for which we report an OR of 1·01 (95% CI 0·98-1·05) or prevalence, for which we report an OR of 0·99 (95% CI 0·96-1·02). IMD is positively associated with prevalence with an OR of 1·03 (95% 1·01-1·06), while the association with test positivity is less definite, with OR of 1·02 (95% CI 0·99-1·05).

### Joint effects of deprivation and ethnicity

Figure 2 shows the modelled average test positivity and debiased prevalence over the entire period of analysis. We observe a similar pattern of results for both outcomes, although prevalence values are on a lower scale. Specifically, the weekly test positivity rate ranges from 2·97% in areas that are mostly White and characterised by low deprivation to 3·35% in the most ethnically diverse and deprived areas, which corresponds to a 13% increase. Similarly, moving from areas with low deprivation and low BAME population to areas with high deprivation and high BAME population, the weekly debiased prevalence shows a 10% increase, changing from 0·37% to 0·41%. The overall effects of IMD and BAME are similar: for instance, among areas with high deprivation, changing the BAME population from low to high changes the test positivity rate from 3·18% to 3·35%; in parallel, among areas with high proportion of BAME population, changing deprivation from low to high changes the test positivity rate from 3·13% to 3·35%.

The effect varies across the different BAME subgroups (Figures 6 and 7 in Supplementary material). Areas where most of the non-White population identifies as Black are characterised by values of test positivity and prevalence similar to mostly White areas, while for areas with a large BAME population that is mostly non-Black we observe an increase in both outcomes relative to mostly White areas.

### Longitudinal variation in effects of deprivation and ethnicity

Figure 3 (top) illustrates the time-varying nature of the effects of BAME and IMD on test positivity. In August-September 2020 there is a 51% increase on the OR scale in test positivity for areas with high deprivation and BAME population (violet line), compared to areas with low deprivation and low BAME (i.e. mostly White) population (red line) and a 30% increase for areas with high deprivation, but relatively small BAME population (pink line). Later, at the peak of the second wave (in December 2020 and January 2021) the effect of BAME becomes more pronounced, reaching a 19% increase in test positivity for areas with large BAME population and low deprivation (light blue line). During the same period there is a less clear gradient across the other profiles. This is expected given the emergence of the Alpha variant and the resulting high disease prevalence across the country at this time, leading to infections spreading more easily across all communities. From February 2021 social deprivation and ethnic composition alternate in which one has a stronger association with test positivity, but with OR generally lower than during the second wave, reaching a peak of 1·23 in May 2021 in the most deprived and most ethnically diverse areas. A similar pattern is seen for debiased prevalence with overall slightly lower OR values than for test positivity.

Finally, the effect profiles of BAME subgroups across the study period (Figure 4), indicate that prior to July 2021 areas with a larger South Asian and Other BAME population were constantly at increased risk for both outcomes (yellow line) relative to areas with predominantly White population. An exception to this is June-July 2020 and March 2021 when areas with a large South Asian population but low in Other BAME show the highest increase (orange line) compared to the reference profile. As the Delta variant was first identified in England in April 2021^7^, this could explain the surge in test positivity among areas with large South Asian populations around that period. There appears to be no substantial difference in test positivity or prevalence between areas with small and large Black population (Figure 8 in the Supplementary material).

## Discussion

Using a flexible spatio-temporal framework, we investigated the effects of social and ethnic inequalities on two LTLA-level measures of SARS-CoV-2 infection load. Overall, we found that area-level percentage of BAME population and area-level deprivation (IMD) are positively associated both with test positivity and with prevalence. When disentangling the effects of ethnicity, we found specifically that areas with a high percentage of South Asian and Other BAME populations were more adversely affected, but not areas with a high percentage of individuals identifying as Black. Moreover, we found that the impact of the two covariates varies over time: deprivation had a stronger effect during the first period of the analysis (until October 2020) whereas the effect of ethnicity became more pronounced during part of the second wave, in December-January (Figure 3). From the spring of 2021 onwards there is a less clear distinction between the impact of BAME and IMD on either outcome. The consistency of results across the two outcome measures leads us to conclude that disease exposure or susceptibility, rather than testing access and habits, differs between areas with those community-level characteristics.

This is the first study to evaluate the impact of social and ethnic inequalities on infections over the whole course of the pandemic so far (excluding the first few months when testing was not widely available at the community level). It is also the first study to model how these effects have evolved over time. Additionally, as we consider test positivity rate and an unbiased estimate of prevalence proportions, we can evaluate the impact of deprivation and ethnicity on both the monitoring and spread of the pandemic, robustly and free from potential confounding by testing ascertainment bias. Lastly, our flexible space time random effects formulation captures the constantly evolving dynamics of the disease arising from emergence of new variants or interventions such as lockdowns, allowing our analyses to automatically adjust for these potential confounders.

A limitation of our study is that the demographic data considered may not fully represent the structure of the population at the time of analysis. For local population sizes, we used mid-year population estimates from 2020 as reference (the most up-to-date available at the time of the study), hence neglecting any changes occurring during the time of the analysis. More critically, we retrieved the proportion of BAME population from the 2011 Census (the most up to date source available at the time of the study). We work under the implicit assumption that the ethnic composition of LTLAs has not changed drastically, but we cannot verify this. Additionally, this study shows an area-level association between inequalities and SARS-CoV-2 infection burden. Given that we consider aggregated data, the analysis may suffer from ecological bias (17) (18).

Our ecological framework is similar to (8) but we focus on SARS-CoV-2 infections rather than mortality and, crucially, adjusts for spatiotemporal dependence in the residuals, a necessary adjustment for obtaining correct estimates of the precision of the effect of deprivation and ethnicity (19). Additionally, while Rose et al. focus on the first wave of the pandemic (8), we consider a longer time period. Our results are consistent with other findings of higher test positivity for BAME groups, but the dynamic nature of our analysis allows us to evaluate how this effect changes during the different phases of the pandemic. Additionally, by considering ethnic composition in conjunction with deprivation, we are able to assess the results for combinations of these two variables, disentangling which variable has a greater impact on test positivity and prevalence at different times. For instance, we show how the nature of the association changed over time, with deprivation being more important in discriminating COVID-19 burden during the start of the second wave of the pandemic and, particularly, in the third national lockdown. This could be a manifestation of financial hardship and/or pandemic fatigue leading to lower adherence to restrictions.

Conversely, in the last few months, we saw the effect of BAME become more prominent, perhaps due to factors like employment in essential sectors and usage of public transport. We found that the effect was not uniform across all BAME groups: the proportion of Black population within an area was not associated with the outcomes as strongly as the proportion of the South Asian and the Other BAME subgroups. Previous reports (1) highlighting an increased test positivity rate for those identifying as Black are limited to the first wave of the pandemic (February - August 2020). Mathur et al. reported that in September - December 2020 the increased test positivity rate in the Black group disappeared (2). Our results are consistent with their findings, even though we only consider Pillar 2 PCR tests while they considered all Pillar 1^8^ and 2 tests combined. In addition, we observe an increased test positivity for South Asians and Other BAME groups for most of the period under analysis prior to July 2021, thus replicating and extending findings reported by Mathur et al. in the first part of the second wave (2).

To conclude, this nationwide population-based study adds to evidence on the impact of social and ethnic inequalities on SARS-CoV-2 infections. Additionally, it shows that, as the pandemic evolved, with the introduction of new variants as well as new policy responses, so did the susceptibility of different communities. We believe that with the constantly changing epidemic dynamics, it is important to continually monitor how different communities are responding, in order to inform relevant policies aimed at eliminating social inequality in COVID-19 burden.

## Ethics statement

This research has been approved by the Alan Turing Institute’s Ethics Advisory Group, nb C2103-105.

## Authors contributions

TP: conceptualisation, methodology, formal analysis, visualisation, writing - original draft

RJ: methodology, formal analysis, writing - original draft

BJ: methodology, formal analysis, writing - review & editing

PD, SR, CH: conceptualisation, methodology, funding acquisition, writing - review and editing

GN: conceptualisation, methodology, formal analysis, writing - review and editing

AM, RK: data curation

MB: conceptualisation, methodology, visualisation, supervision, writing - original draft

## Footnotes

1 https://www.ethnicity-facts-?gures.service.gov.uk/uk-population-by-ethnicity/demographics/people-living-in-deprived-neighbourhoods/latest#data-sources

2 Pillar 2 is the UK government testing programme for the wider population encompassing tests for those showing symptoms and community mass testing schemes (e.g., in schools).

3 https://www.imperial.ac.uk/medicine/research-and-impact/groups/react-study/

4 IMD is provided by the Ministry of Housing, Communities & Local Government, https://www.gov.uk/government/statistics/english-indices-of-deprivation-2019

5 Provided by the Department for Environment, Food & Rural Affairs, https://geoportal.statistics.gov.uk/datasets/rural-urban-classification-2011-of-local-authority-districts-in-england/about

6 https://www.gov.uk/government/news/prime-minister-announces-new-local-covid-alert-levels

7 https://assets.publishing.service.gov.uk/government/uploads/system/uploads/attachment_data/file/991343/Variants_of_Concern_VOC_Technical_Briefing_14.pdf

8 Pillar 1 is the UK government testing program comprising tests conducted in hospitals and care homes.

