## Supplementary Material for "Time varying association between deprivation, ethnicity and SARS-CoV-2 infections in England: a space-time study"

### 1 Model specification

Let $i = 1, \ldots, n = 312$ index the spatial unit (LTLA), $t = 1, \ldots, 68$ index the week and $j = 5-12, \ldots, 65+$ index age groups.

#### 1.1 Test Positivity

Let $B_{itj}$ be the number of Pillar 2 positive tests in LTLA $i$ during week $t$ for age-group $j$ and $n_{itj}$ the total number of processed Pillar 2 tests. We assume $B_{itj}$ follows a Binomial distribution

$$B_{itj}|p_{itj} \sim \text{Bin}(p_{itj}, n_{itj}) \qquad \forall i, j, t.$$

We use the following logistic regression to assess the effect of the variables of interest on the test positivity $p_{itj}$:

$$\begin{aligned}\text{logit}(p_{itj}) = &\beta_0 + \alpha_j + \\ &\beta_1 \texttt{IMD}_i + \beta_2 \texttt{BAME}_i + \\ &\gamma_1 \texttt{Urban}_i + \gamma_2 \texttt{Vax}_i + \\ &\delta_{1,m} \texttt{IMD}_i + \delta_{2,m} \texttt{BAME}_i + \\ &\lambda_i + \omega_t + \xi_{it} \qquad \forall i, j, t.\end{aligned}$$

When considering the distinct BAME subgroups, the model can be reformulated as:

$$\begin{aligned}\text{logit}(p_{itj}) = &\beta_0 + \alpha_j + \\ &\beta_1 \texttt{IMD}_i + \beta_2 \texttt{Black}_i + \beta_3 \texttt{SouthAsian}_i + \beta_4 \texttt{Other}_i \\ &\gamma_1 \texttt{Urbanicity}_i + \gamma_2 \texttt{Vax}_i + \\ &\delta_{1,m} \texttt{IMD}_i + \delta_{2,m} \texttt{Black}_i + \delta_{3,m} \texttt{SouthAsian}_i + \delta_{4,m} \texttt{Other}_i \\ &\lambda_i + \omega_t + \xi_{it} \qquad \forall i, j, t.\end{aligned}$$

The model specification includes a global intercept ($\beta_0$), an age specific intercept ($\alpha_j$), the overall effects of the covariates of interest (captured at the LTLA level), as well as month-specific covariate effects $\delta_{k,m(t)}$, where $m(t)$ represents which month week $t$ belongs to. We impose a Random Walk 1 structure with sum to zero constraints on $\delta_{k,m(t)}$, i.e.

$$\delta_{k,m(t)}|\delta_{k,m(t)-1} \sim \text{Norm}\left(\delta_{k,m(t)-1}, \sigma^2_{\delta_k}\right) \qquad k = 1, \ldots, 4 \quad \forall t.$$

The sum-to-zero constraints ensure that each $\delta_{k,m(t)}$ measures the difference of the effect of month $m(t)$ with respect to the global average $\beta_k$.

Following [5, 4] we model $\boldsymbol{\lambda} = (\lambda_1, \ldots, \lambda_n)$, the random effect accounting for the spatial autocorrelation across LTLAs, as

$$\boldsymbol{\lambda} = \frac{1}{\tau}\left(\sqrt{1-\rho}\boldsymbol{v} + \sqrt{\rho}\boldsymbol{u}\right).$$

$\boldsymbol{u} = (u_1, \ldots, u_n)$ is a spatially structured random effect with prior distribution

$$\boldsymbol{u} \sim \text{Norm}(\boldsymbol{0}, \boldsymbol{Q}^{-})$$

where $\boldsymbol{Q}^{-}$, is the inverse of the precision matrix of a Besag model, scaled in the sense of [6]. $\boldsymbol{v} = (v_1, \ldots, v_n)$ is an i.i.d. Gaussian random effect [1], that is

$$\boldsymbol{v} \sim \text{Norm}(\boldsymbol{0}, \boldsymbol{I})$$

where $\boldsymbol{I}$ is the identity matrix. To account for the time dependence, $\omega_t$ can be modelled through a random walk of order 2, that, given $\Delta^2\omega_t = \omega_t - 2\omega_{t+1} + \omega_{t+2}$, can be formalized as

$$\Delta^2\omega_t|\sigma^2_\omega \sim \text{Norm}\left(0, \sigma^2_\omega\right).$$

Both $\boldsymbol{\lambda}$ and $\boldsymbol{\omega}$ imply some smoothing, hence they help highlight the persistent patterns in the data, but they cannot account for transient anomalies. We thus include a spatio-temporal interaction term $\xi_{it}$ to account for the residual local variability from the general, structured spatio-temporal trend. We assume that

$$\xi_{it} \sim \text{Norm}(0, \sigma^2_\xi)$$

so that $\xi_{it}$ is unstructured in both space and time (or Type I interaction in the sense of [2]). This is critical for highlighting anomalies as the lack of temporal and spatial structure prevents the interaction term to smooth away unexpected but transient behavior.

Finally we assign a Gaussian prior with sum to zero constraint to the parameter $\alpha_j$ for all $j$, we set a minimally informative Gamma$(1, 5 \times 10^{-5})$ on the inverse of $\sigma^2_\delta, \sigma^2_\omega, \sigma^2_\xi$ and a minimally informative Norm$(0, 1000)$ prior for the fixed effect coefficients $\beta_0, \beta_k, \gamma_s$, with $k = 1, \dots, 4$ and $s = 1, 2$.

#### 1.2 Debiased Prevalence

We build on the framework of [3], which allows us to estimate a distribution for the number of cases. Let $I_{itj}$ be the posterior median of such distribution for LTLA $i$, week $t$ and age-group $j$. We assume $I_{itj}$ follows a Binomial distribution

$$I_{itj} | p_{itj} \sim \text{Bin}(p_{itj}, n_{ij}) \qquad \forall i, j, t$$

where $n_{ij}$ the population of LTLA $i$ for age-group $j$ as retrieved by the ONS mid-year population estimates for 2020. Similar to the model for test positivity, we model the debiased prevalence $p_{itj}$ as

$$\begin{aligned}
\text{logit}(p_{itj}) = &\beta_0 + \alpha_j + \\
&\beta_1 \texttt{IMD}_i + \beta_2 \texttt{Black}_i + \beta_3 \texttt{SouthAsian}_i + \beta_4 \texttt{Other}_i \\
&\gamma_1 \texttt{Urbanicity}_i + \gamma_2 \texttt{Vax}_i + \\
&\delta_{1,m} \texttt{IMD}_i + \delta_{2,m} \texttt{Black}_i + \delta_{3,m} \texttt{SouthAsian}_i + \delta_{4,m} \texttt{Other}_i \\
&\lambda_i + \omega_t + \xi_{it} \qquad \forall i, j, t.
\end{aligned}$$

with all model components the same as specified in the previous section.

| | Outcome | Exposures | Confounders |
|---|---|---|---|
| **Main Analysis 1** | PCR Test Positivity | IMD score<br>% BAME population | Age<br>Urbanicity<br>Vaccination Uptake |
| **Main Analysis 2** | Prevalence Estimate | IMD score<br>% BAME population | Age<br>Urbanicity<br>Vaccination Uptake |
| **Main Analysis 3** | PCR Test Positivity | IMD score<br>% Black population<br>% South-Asian population<br>% Other BAME population | Age<br>Urbanicity<br>Vaccination Uptake |
| **Main Analysis 4** | Prevalence Estimate | IMD score<br>% Black population<br>% South-Asian population<br>% Other BAME population | Age<br>Urbanicity<br>Vaccination Uptake |
| **Sensitivity Analysis 1** | PCR and LF Test Positivity | IMD score<br>% Black population<br>% South-Asian population<br>% Other BAME population | Age<br>Urbanicity<br>Vaccination Uptake |
| **Sensitivity Analysis 2** | PCR Test Positivity | IMD score<br>% Black population<br>% South-Asian population<br>% Other BAME population | Age<br>Urbanicity<br>Vaccination Uptake<br>Lockdown |
| **Sensitivity Analysis 3** | PCR Test Positivity | IMD score<br>% Black population<br>% South-Asian population<br>% Other BAME population | Age<br>Urbanicity |

Table 1: Summary of outcomes and covariates included in the all the Analyses (Main and Sensitivity). Random effect specification remains the same as specified in Section 1.

### 2 Additional figures

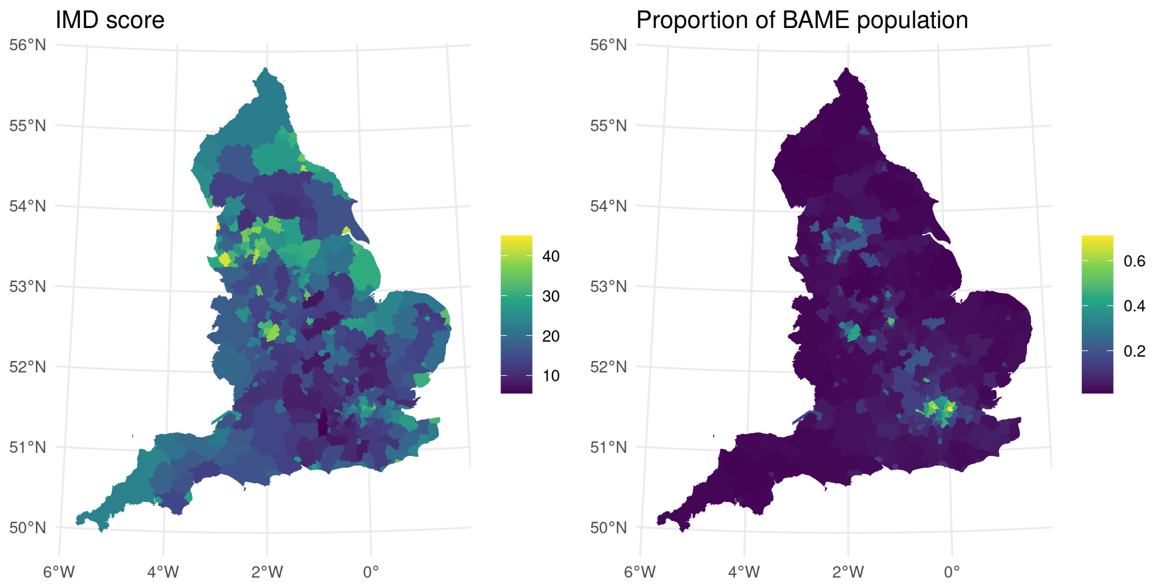


Figure 1: Spatial distribution of the two main exposures, IMD score and proportion of resident BAME population, by LTLA.

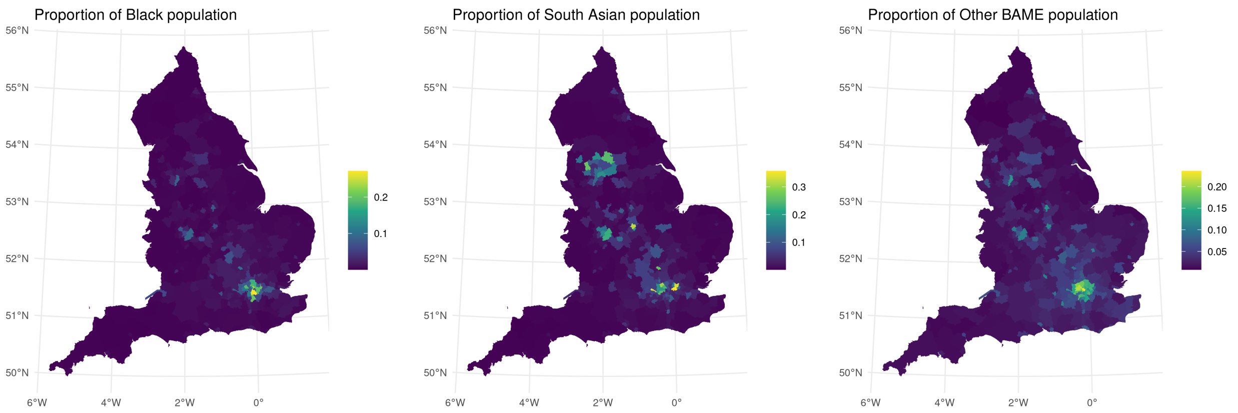


Figure 2: Spatial distribution of the proportion of resident BAME population in the total resident population for the three subgroups considered in the analyses: Black, South Asian and Other BAME.

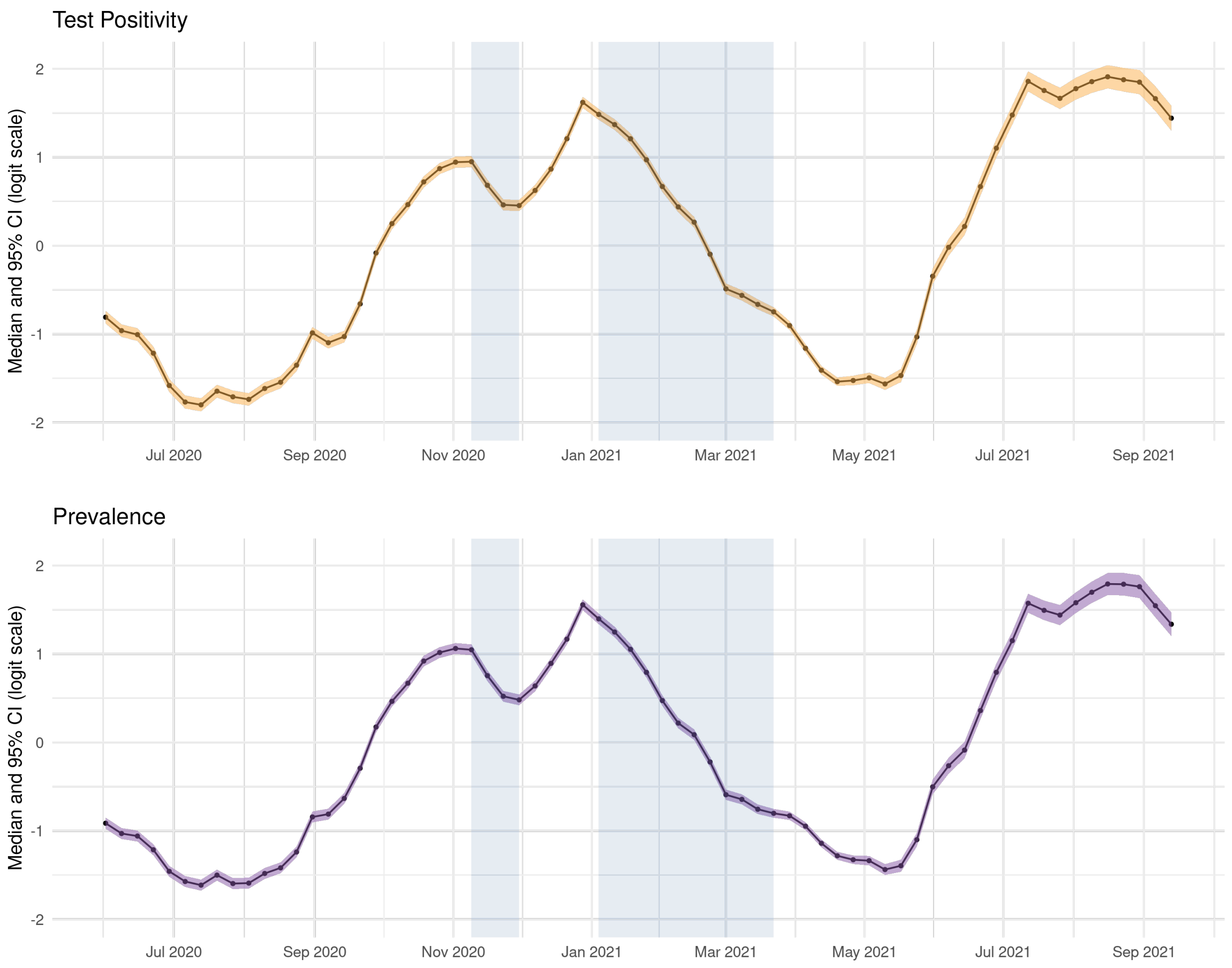


Figure 3: Posterior median and 95% credible intervals (CI) for the temporal trend included in the spatio-temporal model on the logit scale for the main analysis with disaggregated BAME subgroups. Top panel refers to the model with test positivity as outcome, bottom panel to debiased prevalence.

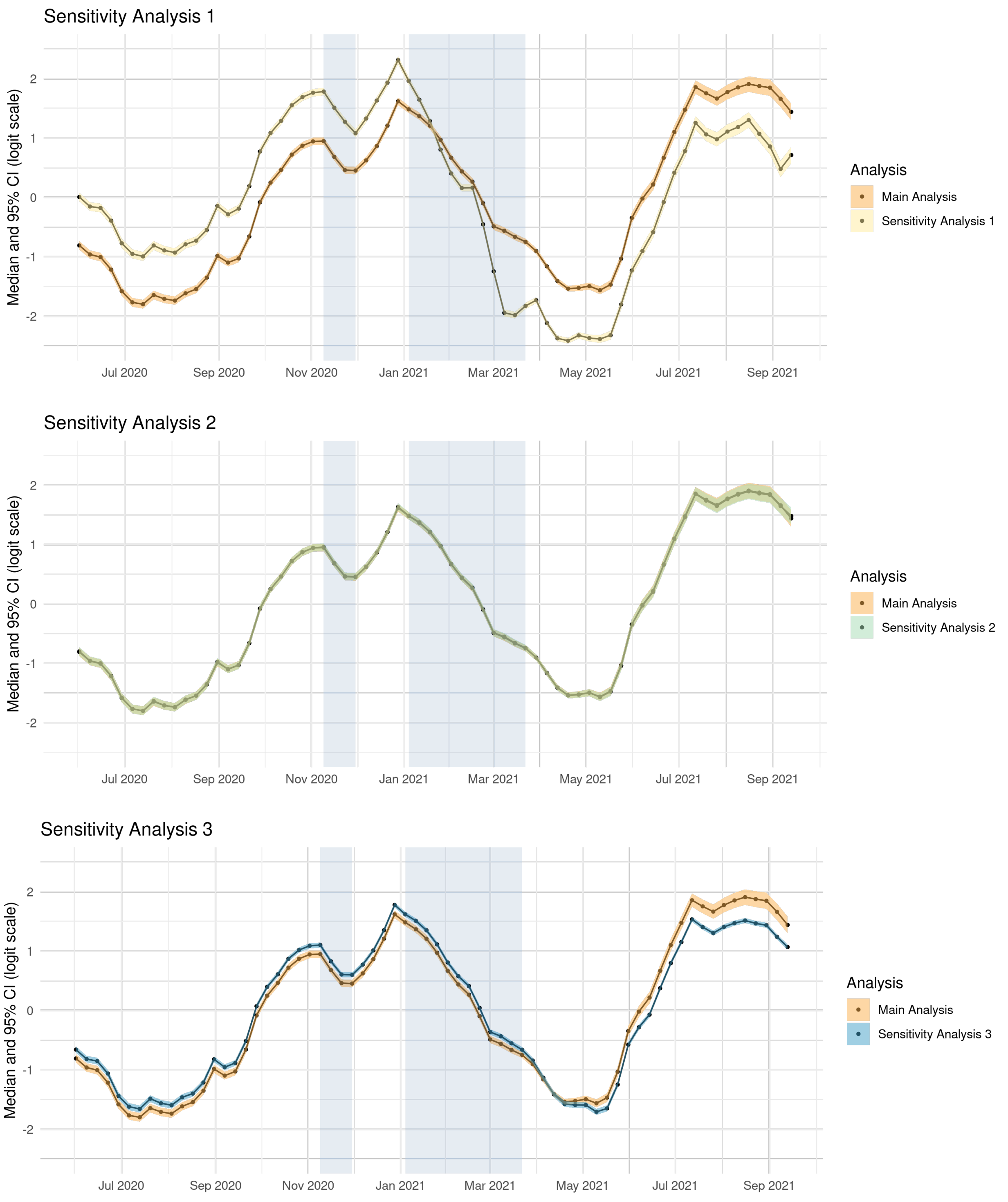


Figure 4: Posterior median and 95% credible intervals (CI) for the temporal trend included in the spatio-temporal model on the logit scale for the main analysis with disaggregated BAME subgroups. Sensitivity analysis 1 (yellow - top panel) uses combined PCR and LF test positivity as outcome measure while sensitivity analyses 2 (green - central panel) and 3 (light blue - bottom panel) use only PCR test positivity. Sensitivity analysis 1 includes IMD, BAME%, age, urbanicity and vaccination as covariates; sensitivity analysis 2 includes lockdown in addition to these covariates; sensitivity analysis 3 removes vaccination from the covariates. When including LFT in the analysis the summer peak is lower than the two winter ones (top panel). This might potentially reflect the change in testing policy: LFT became more widespread as businesses were encouraged to sign up for a testing scheme for their staff, increasing the denominator of the positivity rate. The summer peak is also somewhat less pronounced when removing vaccine uptake from the confounders; in this case the temporal random effect implicitly accounts for the fact that higher vaccination rates lead to a reduction in infections (bottom panel). Finally, the time pattern does not change when we include a lockdown indicator, suggesting that the temporal component already captures much of this effect (central panel).

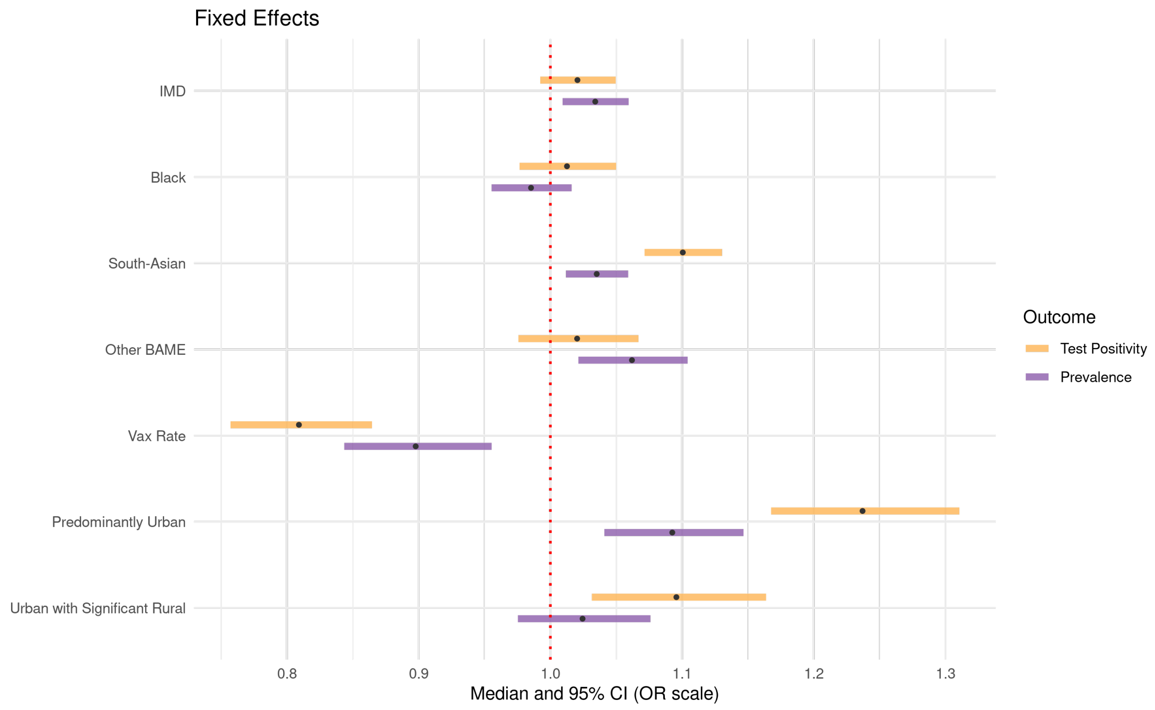


Figure 5: Posterior median (in black) of the fixed effects ($\boldsymbol{\beta}$, $\boldsymbol{\gamma}$) for the main analysis on test positivity and debiased prevalence and corresponding 95% credible intervals (CI). Results are reported on the Odds Ratio (OR) scale.

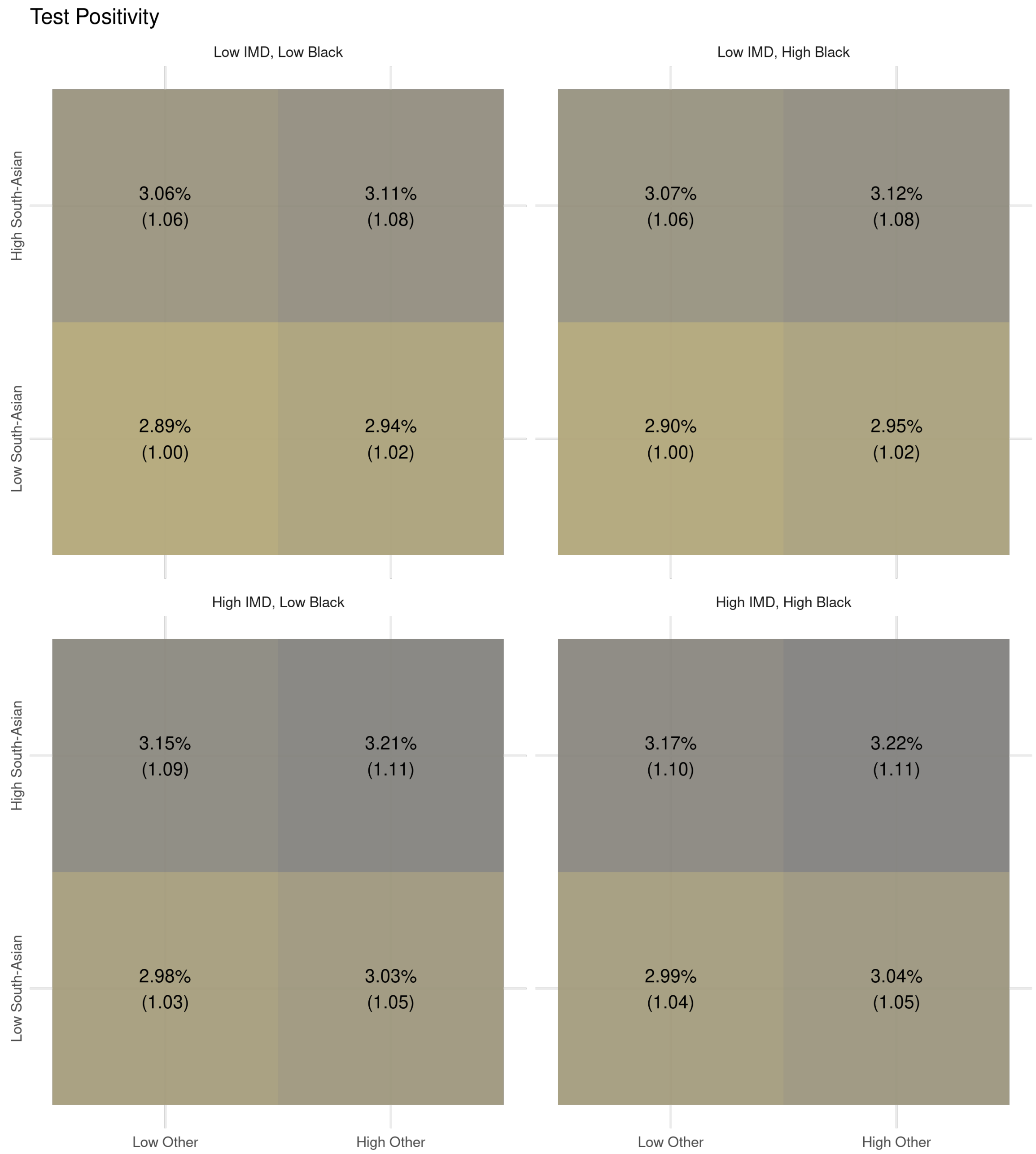


Figure 6: Test positivity for profiles of ethnicity (disaggregated by sub-group) and deprivation. Each tile represents the average weekly test positivity over the entire period of analysis, obtained as output of the model including ethnicity, IMD, confounders and the spatio-temporal correlation structure. In parenthesis we report the relative change in the outcome between each profile and the reference, defined as low deprivation and low Black, South Asian and Other BAME population.

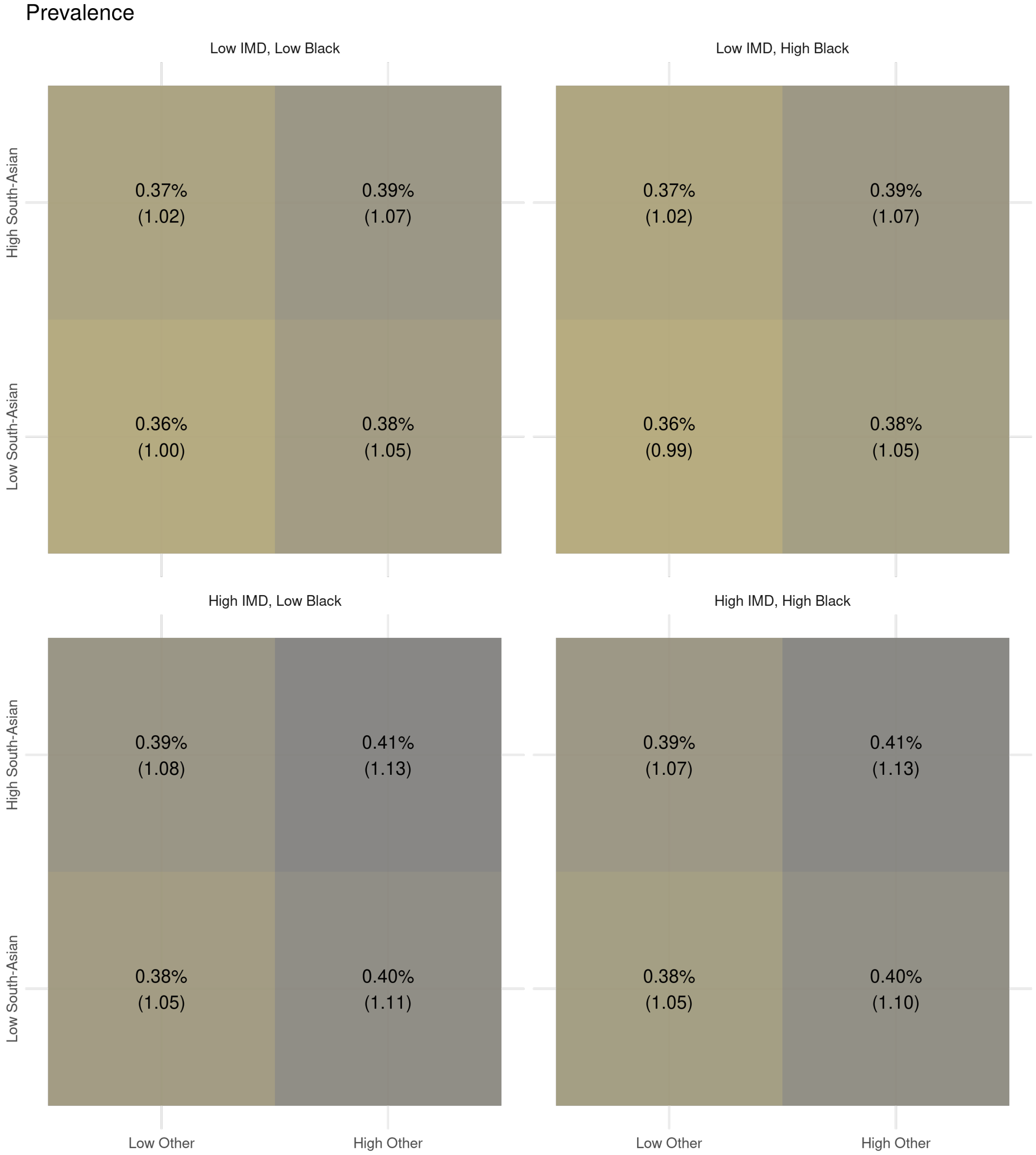


Figure 7: Debiased prevalence for profiles of ethnicity (disaggregated by sub-group) and deprivation. Each tile represents the average weekly debiased prevalence over the entire period of analysis, obtained as output of the model including ethnicity, IMD, confounders and the spatio-temporal correlation structure. In parenthesis we report the relative change in the outcome between each profile and the reference, defined as low deprivation and low Black, South Asian and Other BAME population.

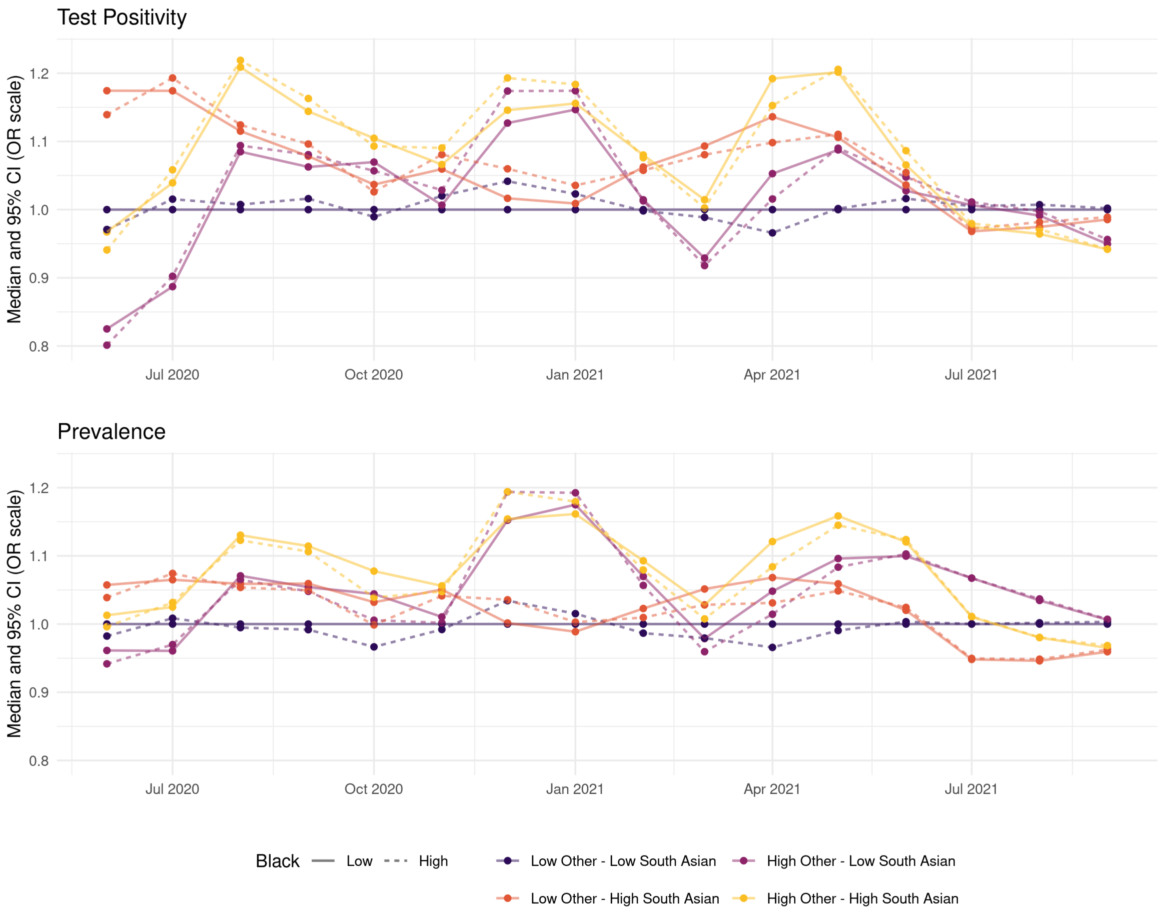


Figure 8: Time-varying test positivity (top) and debiased prevalence (bottom) for profiles of ethnicity disaggregated by BAME subgroups (Black, South Asian, Other). Each line represents the monthly median odds ratio for each profile relative to a population with a low percentage of all BAME subgroups. The mean odds ratio is the output from the model including ethnicity, IMD, confounders and the spatio-temporal correlation structure.

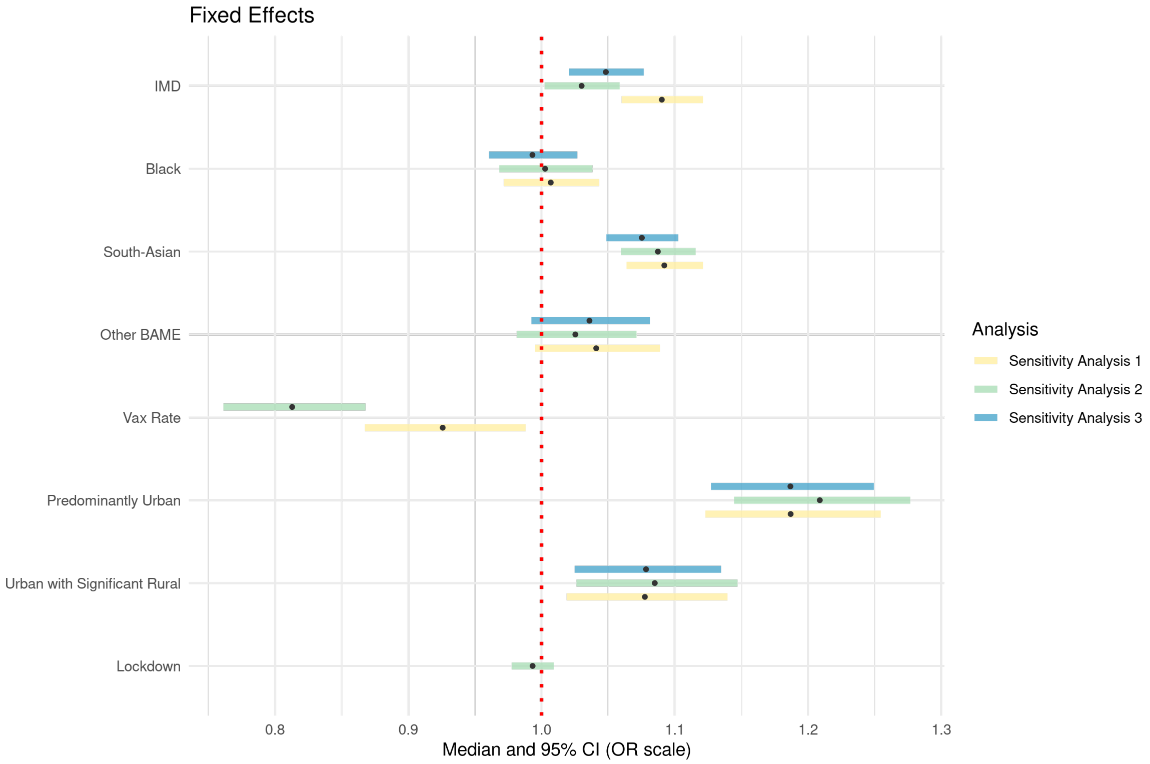


Figure 9: Posterior median (in black) of the fixed effects ($\boldsymbol{\beta}, \boldsymbol{\gamma}$) for the sensitivity analyses and corresponding 95% credible intervals (CI). Results are reported on the Odds Ratio (OR) scale. Sensitivity analysis 1 (yellow) uses combined PCR and LF test positivity as outcome measure while sensitivity analyses 2 (green) and 3 (light blue) use only PCR test positivity. Sensitivity analysis 1 includes IMD, BAME%, age, urbanicity and vaccination as covariates; sensitivity analysis 2 includes lockdown in addition to these covariates; sensitivity analysis 3 removes vaccination from the covariates. The effect of the disaggregated BAME subgroups is robust with respect to the testing strategy and does not change when we include Lateral Flow (LF) tests in the outcome while the effect of IMD is stronger when considering combined PCR and LF test positivity (sensitivity analysis 1).

[3] George Nicholson, Brieuc Lehmann, Tullia Padellini, Koen B Pouwels, Radka Jersakova, James Lomax, Ruairidh E King, Ann-Marie Mallon, Peter J Diggle, Sylvia Richardson, Marta Blangiardo, and Chris Holmes. Local prevalence of transmissible sars-cov-2 infection: an integrative causal model for debiasing fine-scale targeted testing data. *medRxiv*, 2021.

[4] Andrea Riebler, Sigrunn H Sørbye, Daniel Simpson, and Håvard Rue. An intuitive bayesian spatial model for disease mapping that accounts for scaling. *Statistical methods in medical research*, 25(4):1145–1165, 2016.

[5] Daniel Simpson, Håvard Rue, Andrea Riebler, Thiago G Martins, Sigrunn H Sørbye, et al. Penalising model component complexity: A principled, practical approach to constructing priors. *Statistical science*, 32(1):1–28, 2017.

[6] Sigrunn Holbek Sørbye and Håvard Rue. Scaling intrinsic gaussian markov random field priors in spatial modelling. *Spatial Statistics*, 8:39–51, 2014.
